# Uptake and factors associated with cervical cancer screening among women aged 18-49 years at a public hospital in Coastal Kenya

**DOI:** 10.1101/2025.06.24.25330178

**Authors:** Dennis Mose, Geoffrey Katana, Stevenson K. Chea, Phillip Ayieko

**Author notes:** Corresponding author (SC). These authors contributed equally to this work (Stevenson K. Chea, Phillip Ayieko).

## Abstract

Cervical cancer remains a public health concern in sub-Saharan Africa. In Kenya, data on cervical cancer screening is limited therefore hindering planning of targeted interventions. We aimed to determine the prevalence and factors associated with cervical cancer screening among women aged 18 - 49 years at a public hospital in Coastal Kenya.

A cross-sectional design was used. Women attending outpatient departments were recruited using systematic random sampling and an interviewer-administered questionnaire administered (n = 315). Proportion of women reporting cervical cancer screening uptake was determined. Logistic regression was used to examine associations between cervical cancer screening uptake and sociodemographic characteristics.

Overall, 56 (17.8% [95% CI: 13.7 – 22.5]) participants reported to have been screened for cervical cancer. Factors associated with cervical cancer screening uptake included older age (adjusted odds ratios, [95% CI], p – value: 2.3 [1.0 – 5.0], p = 0.034), secondary/tertiary level of education (3.3 [1.6 – 6.5], p < 0.001) and history of sexually transmitted infection (STI) (2.4 [1.2 – 4.7], p = 0.009).

Uptake of cervical cancer screening was low. Intensifying education campaigns on cervical cancer screening especially among women who are young, uneducated and with no history of STI may help scale up cervical cancer screening uptake.

## Introduction

Though preventable, cervical cancer is regarded as the fourth most common cancer in women globally and a major public health concern in low and medium income countries (LMICs) [1–4]. About, 85% of cervical cancer burden occurs in Sub-Saharan Africa (sSA) [5, 6], contributing to approximately 20.8% of all cancers in women and 14.2% of all cancer-related deaths in women [7]. The prevalence of cervical cancer in LMICs is two-fold higher and its death rates three times as high as those in high-income countries [8]. The human papillomavirus (HPV), an infection commonly acquired through sexual intercourse, is the main cause of cervical pre-cancer and cancer cases, accounting for 99% of cases [9]. Importantly, HPV is associated with other preventable sexually transmitted infections including human immunodeficiency virus (HIV) [10] with studies revealing the connection between HIV and the incidence of cervical cancer [11].

Recent health promotion efforts have shown that one-third of cervical cancer cases can be controlled by early detection through screening and prompt treatment [12]. Early detection of cervical cancer increases the chance of effective intervention through targeting behavioral and individual factors. The World Health Organization recommends cervical cancer screening for all women aged 30-49 years [13].

Some of the factors that have been shown to be associated with uptake of cervical cancer screening include age and level of education. Previous studies from Kenya have estimated the uptake of cervical cancer screening to be in the range of 1-36% [14–16]. However, none of these studies was conducted at the site of the current study. Altogether, efforts on cervical screening have been done in Kilifi county with limited documentation regarding the prevalence of cervical cancer screening therefore hindering planning of targeted interventions. We aimed at determining the prevalence and correlates of cervical cancer screening among women aged 18 - 49 years attending care at Malindi sub County hospital in Kilifi, along the Kenyan Coast.

## Methods

### Study site

The study was conducted at Malindi Sub county hospital. The hospital serves as a referral facility to the neighboring Counties with a catchment population of 287,670 people out of which 23.2% (66,739) are women of reproductive age [17, 18]. Cervical cancer screening has been integrated with HIV/other sexually transmitted infections (STI) care and is offered in Maternal child health and family planning clinic (MCH/FP), comprehensive care clinic (CCC) daily and weekly in general out-patient (OPD) departments in the hospital. An average of 20 women are screened weekly using Visual Inspection with Acetic Acid (VIA)/Visual Inspection using Lugol’s Iodine (VILLI) approach [19, 20]. The services are offered mostly by trained nurses and doctors. There is occasional outreach and in reach activities conducted by the facility in collaboration with other implementing partners. The hospital uses screen and treat approach where those with positive precancerous lesions are treated with cryotherapy machine. Those with cancerous tumors are referred to gynecology clinic for further management and linked for long term care in the regional referral hospital.

### Study design

A cross-sectional design was used. Women attending CCC, MCH/FP and OPD between 25^th^ February 2015 and 13^th^ April 2015 were recruited and interviewed using an interviewer-administered questionnaire (n = 315). Women aged between 18 and 49 years who had not undergone hysterectomy were eligible.

### Sampling strategy

A systematic random sampling strategy was used. Based on enquiries made to staff in the CCC, MCH/FP and general OPD regarding patient flow, we estimated that the population of women aged 18-49 years attending care in these departments would be above 6000. With an estimated sample size of 328, a sampling interval of 19 was deemed appropriate. As a standard procedure, women presenting for consultation at the hospital CCC, MCH/FP and general OPD department have their details captured in the daily attendance register by a clerk in the order of their arrival. This helped minimize selection bias as the women details were not captured in any order of preference. One clerk is stationed at the reception of each of the three departments. Each clerk was requested to refer every 19^th^ woman, starting from the first woman to arrive, to a research assistant for screening and enrollment if they were eligible and gave consent. A research assistant was stationed in each of the three departments. If the woman did not meet the eligibility criteria or refused to consent, then the 20^th^ one was interviewed. Participants were recruited proportionately from the CCC, MCH/FP and general OPD departments. Assuming a 14.4% prevalence [21] for cervical cancer screening coverage and adjusting for 10% non-response, we estimated that a sample of 328 participants with a 4% precision would be sufficient [22].

### Sources of data

An interviewer-administered questionnaire was used to collect data including sociodemographic characteristics and cervical cancer screening practices. Sociodemographic data included age, marital status, residence and occupation. Questionnaires were checked during data collection for completeness. Participants were interviewed individually in a separate room within the hospital where this took approximately 30-40 minutes. Data collection started on 25^th^ February 2015 and ended on 13^th^ April 2015. Data capture was done in Microsoft Access version 2007.

### Data analysis

Socio-demographic characteristics were summarized using frequencies and percentages. The uptake of cervical cancer screening was determined as the proportion of women who reported to have ever been screened for cervical cancer. Binomial exact 95% confidence intervals (CI) were presented for proportion of women reporting screening uptake. To assess factors associated with uptake of cervical cancer screening, first a univariable logistic regression analysis was done to assess crude associations between each exposure variable and the outcome. Exposure variables with a p-value <0.2 were carried forward to the multivariable model. Decision regarding which exposure variables will be included in the multivariable model may depend on several factors including clinical and statistical significance [23]. In this study we used statistical significance to determine the exposures to carry forward since there was no primary exposure of interest. Literature suggests that a p value of 0.2 or above could be used as a cut off to select exposures to include in final model [23]. Lastly, a multivariable logistic regression model was fitted using a step wise model building approach [24] to assess independent predictors of cervical cancer screening uptake. All analysis was performed in Stata 15.0 (StataCorp. 2017. Stata Statistical Software: Release 15. College Station, TX: StataCorp LLC. 2019) and graphs generated using GraphPad Prism version 8.0.2 (GraphPad software, California).

### Ethical considerations

Ethical approval for this study was granted by the Pwani University Ethics Review Committee (Reference No. ERC/MSc/032/2014). In addition, administrative approval was granted by the Research committee of Kilifi County Government (REF: DOH/KLF/RESCH/VOL.1/17). Written informed consent was obtained from all study participants.

## Results

### Characteristics of participants

Of the 315 participants included in the analysis, majority were 26 – 49 years old (198 [62.9%]), educated up to primary school level or had no formal education (222 [70.5%]), Christians (229 [72.7%]), employed or in business (181 [57.5%]) and married (227 [72.1%]). Further, majority had given birth to at least one child in their life (271 [86.0%]), had never used oral contraceptives (224 [71.1%]), and were not currently smoking and had never smoked in their life time (302 [95.9%]). A further majority had their sexual debut before their 18^th^ birth day (194 [61.6%]), had more than one life time sexual partner (194 [61.6%]) and had never had a sexually transmitted infection (217 [68.9%]) (Table 1).

**Table 1:** Socio-demographic characteristics of women aged 18-49 years attending care at a sub-county public health facility in Coastal Kenya (N=315)

| Characteristic | Category | Urban residence<br>(n = 172)<br>N [%] | Rural residence<br>(n = 143)<br>N [%] | Overall<br>(N=315)<br>N [%] |
| --- | --- | --- | --- | --- |
| Age in years Mean [26] | - | 28.9 [6.9] | 29.5 [7.7] | 29.2 [7.3] |
| Age group [Years] | 18 – 25 | 64 [37.2] | 53 [37.1] | 117 [37.1] |
|  | 26 - 49 | 108 [62.8] | 90 [62.9] | 198 [62.9] |
| Education level | No formal education/primary | 113 [65.7] | 109 [76.2] | 222 [70.5] |
|  | Secondary/tertiary | 59 [34.3] | 34 [23.8] | 93 [29.5] |
| Religion | Christian | 117 [68.0] | 112 [78.3] | 229 [72.7] |
|  | Muslim/other | 55 [32.0] | 31 [21.7] | 86 [27.3] |
| Occupation | Employed/business | 90 [52.3] | 91 [63.6] | 181 [57.5] |
|  | Unemployed | 82 [47.7] | 52 [36.4] | 134 [42.5] |
| Marital status | Married | 124 [72.1] | 103 [72.0] | 227 [72.1] |
|  | Single/widowed/separated/divorced | 48 [27.9] | 40 [28.0] | 88 [27.9] |
| Children ever given birth to | None | 25 [14.5] | 19 [13.3] | 44 [14.0] |
|  | At least one | 147 [85.5] | 124 [86.7] | 271 [86.0] |
| Oral contraceptive use ever | Used | 55 [32.0] | 36 [25.2] | 91 [28.9] |
|  | Never used | 117 [68.0] | 107 [74.8] | 224 [71.1] |
| Duration of contraceptive use<sup>a</sup> | Less than 5 years | 49 [89.1] | 30 [83.3] | 79 [86.8] |
|  | More than 5 years | 6 [10.9] | 6 [16.7] | 12 [13.2] |
| Ever or currently smoking | Yes | 7 [4.1] | 6 [4.2] | 13 [4.1] |
|  | No | 165 [95.9] | 137 [95.8] | 302 [95.9] |
| Age at sex debut [Years] | Above 18 | 75 [43.6] | 46 [32.2] | 121 [38.4] |
|  | 18 and below | 97 [56.4] | 97 [67.8] | 194 [61.6] |
| Lifetime sexual partners | One or none | 58 [33.7] | 63 [44.1] | 121 [38.4] |
|  | More than one | 114 [66.3] | 80 [55.9] | 194 [61.6] |
| Ever had STI<sup>b</sup> | Yes | 46 [26.7] | 52 [36.4] | 98 [31.1] |
|  | No | 126 [73.3] | 91 [63.6] | 217 [68.9] |

<sup>c</sup>Computed from those who had ever used contraceptives [n = 91]
<sup>b</sup>Sexually transmitted infection

### Prevalence of cervical cancer screening uptake and associated factors

Of the 315 participants included in the analysis, 56 (17.8% [95% CI: 13.7 – 22.5]) reported to have been screened for cervical cancer. In univariable analysis, age, education status, religion and ever reporting an STI were significantly associated with the outcome (Table 2). In the multivariable model, age, education status and ever reporting an STI were independent predictors of cervical cancer screening uptake. Education status and ever reporting an STI attenuated religion towards the null. Women aged 26 – 49 years had increased odds of reporting to have been screened for cervical cancer compared to those aged 18 – 25 years (adjusted odds ratios, aOR [95% CI]: 2.3 [1.0 – 5.0], p = 0.034). Equally, women with a secondary/tertiary education had increased odds of reporting to have been screened for cervical cancer compared to those with primary/no formal education (adjusted odds ratios, aOR [95% CI]: 3.3 [1.6 – 6.5], p < 0.001). Lastly, women reporting to have ever had symptoms of STI had increased odds of reporting to have been screened for cervical cancer compared to those who had never had STI symptoms (adjusted odds ratios, aOR [95% CI]: 2.4 [1.2 – 4.7], p = 0.009) (Table 2) (Fig 1).

**Table 2:** Factors associated with uptake of cervical cancer screening among women aged 18-49 years attending care at a sub-county public health facility in Coastal Kenya (N=315).

| Predictor | Category | Screened for<br>cervical<br>cancer n [%] | Not screened<br>for cervical<br>cancer n [%] | Crude OR<br>[95% CI] | p-value | Adjusted OR<br>[95% CI] | p-value |
| --- | --- | --- | --- | --- | --- | --- | --- |
| Age [Years] | 18 - 25 | 10 [8.5] | 107 [91.5] | Ref | Ref | Ref | Ref |
|  | 26 - 49 | 46 [23.2] | 152 [76.8] | 3.2 [1.5 – 6.7] | <0.001 | 2.3 [1.0 – 5.0] | 0.034 |
| Residence | Rural | 24 [16.8] | 119 [83.2] | Ref | Ref |  |  |
|  | Urban | 32 [18.6] | 140 [81.4] | 1.1 [0.6 – 2.0] | 0.674 |  |  |
| Education level | No formal education/primary | 29 [13.1] | 193 [86.9] | Ref | Ref | Ref | Ref |
|  | Secondary/tertiary | 27 [29.0] | 66 [71.0] | 2.7 [1.5 – 4.9] | 0.001 | 3.3 [1.6 – 6.5] | <0.001 |
| Religion | Muslim/African traditional | 9 [10.5] | 77 [89.5] | Ref | Ref | Ref | Ref |
|  | Christian | 47 [20.5] | 182 [79.5] | 2.2 [1.0 – 4.7] | 0.041 | 2.2 [1.0 – 5.1] | 0.047 |
| Occupation | Housewife/student/unemployed | 21 [15.7] | 113 [84.3] | Ref | Ref |  |  |
|  | Farmer/business/employed | 35 [19.3] | 146 [80.7] | 1.2 [0.7 – 2.3] | 0.401 |  |  |
| Marital status | Married | 44 [19.4] | 183 [80.6] | Ref | Ref |  |  |
|  | Single/widowed/separated/divorced | 12 [13.6] | 76 [86.4] | 0.6 [0.3 – 1.3] | 0.234 |  |  |
| Children ever given birth to | None | 3 [6.8] | 41 [93.2] | Ref | Ref | Ref | Ref |
|  | At least one | 53 [19.6] | 218 [80.4] | 3.3 [0.9 – 11.1] | 0.052 | 3.0 [0.8 – 11.4] | 0.095 |
| Oral contraceptive use ever | No | 31 [13.8] | 193 [86.2] | Ref | Ref | Ref | Ref |
|  | Yes | 25 [27.5] | 66 [72.5] | 2.3 [1.2 – 4.2] | 0.005 | 1.7 [0.9 – 3.3] | 0.096 |
| Ever smoked | No | 53 [17.5] | 249 [82.5] | Ref | Ref |  |  |
|  | Yes | 3 [23.1] | 10 [76.9] | 1.4 [0.3 – 5.2] | 0.611 |  |  |
| Age at sex debut | 18 years or below | 36 [18.6] | 158 [81.4] | 1.1 [0.6 – 2.0] | 0.647 |  |  |
|  | Above 18 years | 20 [16.5] | 101 [83.5] | Ref | Ref |  |  |
| Number of sex partners | One or none | 17 [14.1] | 104 [85.9] | Ref | Ref | Ref | Ref |
|  | More than one | 39 [20.1] | 155 [79.9] | 1.5 [0.8 – 2.8] | 0.174 | 1.3 [0.6 – 2.6] | 0.404 |
| Ever had STI | No | 28 [12.9] | 189 [87.1] | Ref | Ref | Ref | Ref |
|  | Yes | 28 [28.6] | 70 [71.4] | 2.7 [1.4 – 4.8] | 0.001 | 2.4 [1.2 – 4.7] | 0.009 |

STI, Sexually transmitted infection; OR, Odds ratio
Note: Only variables with p value <0.2 in univariable analysis were included in final model.

**Figure 1:**
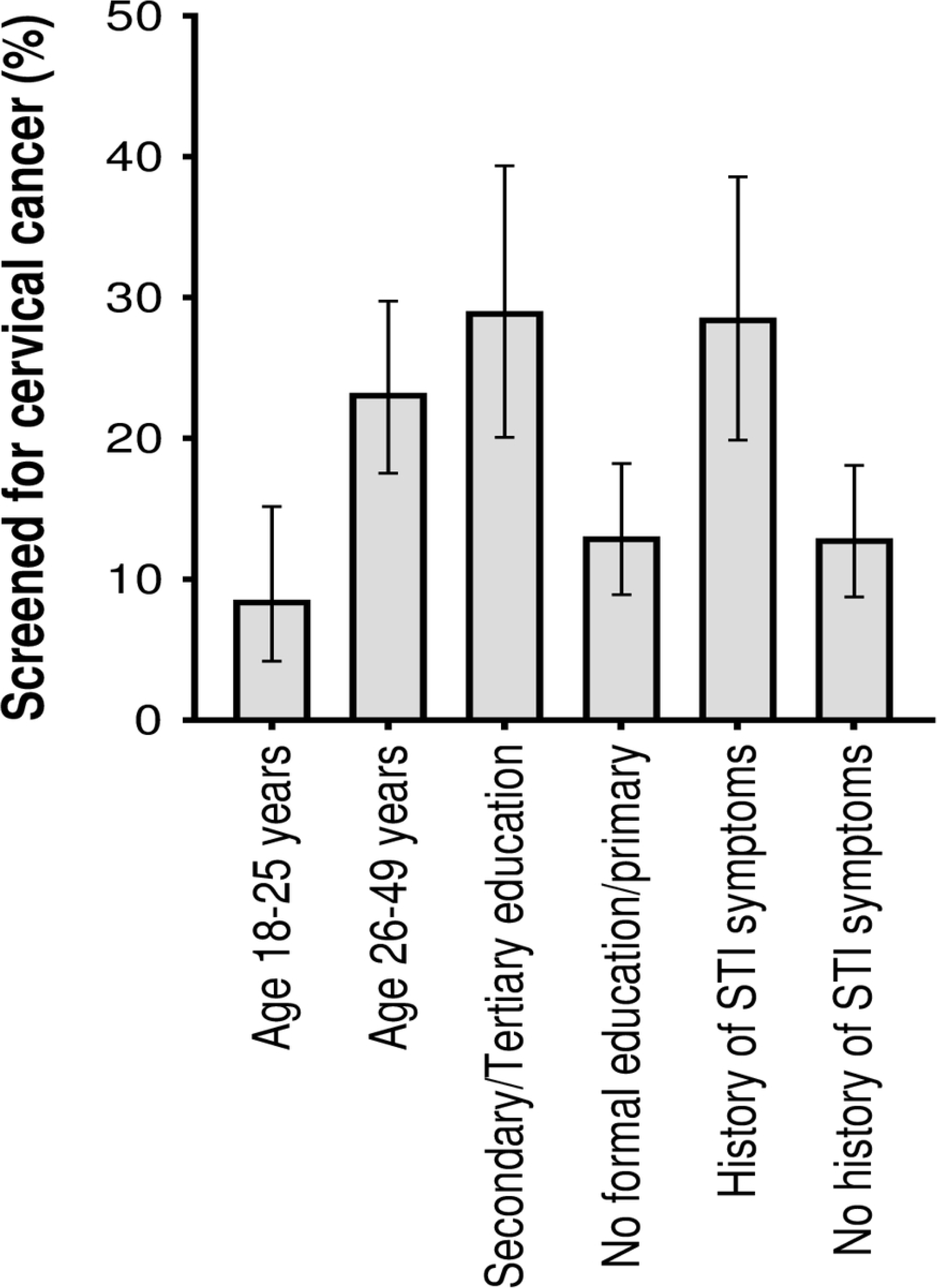
Distribution of factors that showed a significant association with the outcome in multivariable model by cervical cancer screening status (n = 315) STI, sexually transmitted infections

## Discussion

In this study, about one in every five women aged 18 – 49 years reported to have been screened for cervical cancer. Age, education status and ever reporting an STI were independent predictors of cervical cancer screening uptake.

Approximately one in every five women had been screened for cervical cancer which is comparable to the 13.6-18.5% range reported in a systematic review of studies conducted in Ethiopia [25]. Our estimate is also comparable to the 16.4% reported in a nationally-representative survey involving women age 30-49 years in Kenya [16]. A recent Kenyan study that used data of women aged 15-49 years from the 2022 Kenya demographic and health survey reported 16.8% cervical cancer screening uptake which is equally comparable to our estimate [17]. Our estimate is however much lower compared to the 30% from a South African study that enrolled women attending an outreach [26]. Another South African study conducted among rural women aged 18 – 65 years reported 66.8% cervical cancer screening uptake which is equally higher compared to our estimate [27]. The differences between our estimate and those from South Africa could be due to differences in sociodemographic and socio-economic status of the study participants. Importantly, our estimate is much lower compared to the targeted 70% as recommended by WHO [28] and the Kenyan cervical cancer program 2011-2020 [14]. The current study was conducted at a health facility in Kilifi County located in Coastal Kenya. Kilifi county is one of the poorest counties in Kenya with a food poverty rate of 39.3-52.7% [29]. Further, the larger administrative area known as Kilifi County in Kenya mainly comprises of people with low levels of education [29]. Indeed, over 70% of our study participants had primary level of education or no formal education at all. It is possible that financial constraints including lack of transport to the health facility for cervical cancer screening as well as failure to understand the value of screening as a result of low education status may have contributed to the low cervical cancer screening uptake in this population. Further, despite efforts to scale up cervical cancer screening in Kenya, some challenges persist including shortage of clinical staff in health facilities leading to long waiting time [30]. Although these factors were not explored in the current study, they may potentially explain the low uptake of cervical cancer screening in this population. Further, cultural factors were not explored in this study. Previous studies have reported cultural influences on health seeking behaviour including uptake of cervical cancer screening [31]. Future studies may employ a mixed methods approach in order to explore the cultural influences on cervical cancer screening uptake. Notably, uptake of cervical cancer screening among women aged 18-25 years was lower compared to women aged 26-49 years (8.5% *vs* 23.2%, p = 0.001). According to the Kenya national cervical cancer screening guidelines, the primary target for cervical cancer screening are women aged 25-49 years [32]. Women under 25 years of age are screened only if they are at a higher risk of disease. This could explain the discrepancy in the uptake of cervical cancer screening between younger and older women. Although Kenya has established a population-based cervical cancer screening program [33], our findings of low cervical cancer screening uptake imply that this program needs to be strengthened if the WHO targets of elimination of cervical cancer are to be achieved [28]. One way to strengthen this program is to scale up efforts on educating the population about cervical cancer screening and its importance. A recent systematic review has shown that educational interventions are effective in promoting uptake of cervical cancer screening among women [34].

Older women had increased odds of reporting to have been screened for cervical cancer compared to younger women. A multi-site study from Africa [35] and a previous study from Kenya [15] equally found that older age was associated with increased odds of cervical cancer screening. It is plausible that older women have had more experiences in life including more opportunities to learn and appreciate the value of cervical cancer screening hence more likely to be receptive to screening. This implies that efforts to scale up cervical cancer screening uptake in Kilifi County, Coastal Kenya need to include support for younger women. Specifically, such support may involve intensifying education campaigns on cervical cancer screening among young women in this population [34]. Further, it may be important to consider targeting even women under 25 years for screening in order to increase screening uptake in younger women.

Women with a secondary/tertiary education had increased odds of reporting to have been screened for cervical cancer compared to those with primary/no formal education. Previous studies from Ethiopia [25, 36] and South Africa [37] equally found that educated women were more likely to have been screened for cervical cancer. Educated women can have a better awareness about cervical cancer and its screening [37], mostly have decision making power [35], and better health care seeking initiative than non-educated which in turn can result in the utilization of the screening service. This implies that efforts to scale up cervical cancer screening uptake in Kilifi County, Coastal Kenya need to include support for women who are not educated. Such support may include measures to ensure girls remain in school. This will enhance their knowledge and understanding thus be able to adopt good health seeking behaviours including getting screened for cervical cancer.

Women reporting to have ever had symptoms of STI had increased odds of reporting to have been screened for cervical cancer compared to those who had never had STI symptoms. A previous study from Ethiopia also found that women with history of STI had increased odds of reporting to have been screened for cervical cancer [25]. It is possible that women with history of STI symptoms may have had contact with the health service for STI care. It is possible that they received counselling and education on STIs and the importance of screening as per national guidelines [38]. This may have motivated them to seek screening services. The Kenyan government has integrated cervical cancer screening in HIV/STI care [19]. Our finding that women with history of STI are more likely to have been screened possibly implies that this integration is working as it provides a one-stop shop where women attending HIV/STI care are provided with cervical cancer screening services.

One limitation of our study is that participants were recruited from one health facility only. This introduces potential bias and could limit generalizability of our findings to all women in Coastal Kenya and the whole country at large since women attending care in the health facility could be different from those in the community. In addition, cervical cancer screening uptake data was self-reported which introduces potential social desirability bias. Future studies may combine self-reported data with clinical records or observational data. Secondly, the study mainly assessed sociodemographic factors. Future studies may focus on a wide range of factors including health system factors which may have a bearing on uptake of cervical cancer screening. Thirdly, our study utilizes data collected in 2015 which may be considered old. However, our 17.8% prevalence estimate of cervical cancer screening uptake is comparable to the 16.8% reported in a 2024 study that utilized a national representative sample of women of reproductive age from the 2022 Kenya demographic and health survey [17]. The similarity between the two estimates can be taken to mean that the 2015 data to some extent still reflects the current status of cervical cancer screening in Kenya. However, Kenya has since adopted a community based cervical cancer screening approach where eligible women are screened in the community [39]. Unlike the previous facility-based approach, this new approach is expected to scale up the uptake of cervical cancer screening in Kenya once it is fully implemented.

In conclusion, the uptake of cervical cancer screening was low in this population with levels that are substantially lower than international targets for cervical cancer screening. Older women as well as the educated and those with history of STI had higher odds of reporting to have been screened. Intensifying education campaigns on cervical cancer screening with a particular focus on women who are young, uneducated and with no history of STI may help scale up cervical cancer screening uptake in this population.

## Data Availability

Data relating to this study is freely available on figshare 10.6084/m9.figshare.25515388

## Acknowledgements

We acknowledge Prof. Jameela Hassanali of Pwani University for her support throughout the project. Further, we acknowledge all study participants for voluntarily taking part in this study.

## References

1. Cecilia NC, Rosliza AM, Suriani I. Global burden of cervical cancer: A literature review. International Journal of Public Health and Clinical Sciences2017. p. 10–8.

2. Arbyn M, Weiderpass E, Bruni L, de Sanjosé S, Saraiya M, Ferlay J, et al. Estimates of incidence and mortality of cervical cancer in 2018: a worldwide analysis. Lancet Glob Health. 2020;8(2):e191–e203. 10.1016/s2214-109x(19)30482-6 PMC7025157

3. Bray F, Ferlay J, Soerjomataram I, Siegel RL, Torre LA, Jemal A. Global cancer statistics 2018: GLOBOCAN estimates of incidence and mortality worldwide for 36 cancers in 185 countries. CA Cancer J Clin. 2018;68(6):394–424. 10.3322/caac.21492

4. Kafuruki L, Rambau PF, Massinde A, Masalu N. Prevalence and predictors of Cervical Intraepithelial Neoplasia among HIV infected women at Bugando Medical Centre, Mwanza-Tanzania. Infect Agent Cancer. 2013;8(1):45. 10.1186/1750-9378-8-45 PMC3833176

5. Ferlay J, Soerjomataram I, Dikshit R, Eser S, Mathers C, Rebelo M, et al. Cancer incidence and mortality worldwide: sources, methods and major patterns in GLOBOCAN 2012. Int J Cancer. 2015;136(5):E359–86. 10.1002/ijc.29210

6. Kabalika C, Mulenga D, Mazaba ML, Siziya S. Acceptance of Cervical Cancer Screening and its Correlates Among Women of a Peri-Urban High-Density Residential Area in Ndola, Zambia. Int J MCH AIDS. 2018;7(1):17–27. 10.21106/ijma.223 PMC6168797

7. World Health Organization IAfRoC. IARC handbooks of cancer prevention France: International Agency for Research on Cancer; 2005 [Available from: https://zora.onko-i.si/fileadmin/user_upload/publikacije/tuje_publikacije/C14%20-%20IARC%20cervical%20cancer%20screening.pdf.

8. Zhang S, Xu H, Zhang L, Qiao Y. Cervical cancer: Epidemiology, risk factors and screening. Chin J Cancer Res. 2020;32(6):720–8. 10.21147/j.issn.1000-9604.2020.06.05 PMC7797226

9. Boda D, Docea AO, Calina D, Ilie MA, Caruntu C, Zurac S, et al. Human papilloma virus: Apprehending the link with carcinogenesis and unveiling new research avenues (Review). Int J Oncol. 2018;52(3):637–55. 10.3892/ijo.2018.4256 PMC5807043 journal, but had no personal involvement in the reviewing process, or any influence in terms of adjudicating on the final decision, for this article.

10. Wanyenze RK, Bwanika JB, Beyeza-Kashesya J, Mugerwa S, Arinaitwe J, Matovu JKB, et al. Uptake and correlates of cervical cancer screening among HIV-infected women attending HIV care in Uganda. Glob Health Action. 2017;10(1):1380361. 10.1080/16549716.2017.1380361 PMC5678455

11. Gichangi P, De Vuyst H, Estambale B, Rogo K, Bwayo J, Temmerman M. HIV and cervical cancer in Kenya. Int J Gynaecol Obstet. 2002;76(1):55–63. 10.1016/s0020-7292(01)00560-4

12. Abenwie SN EM, Edo’o VD, Hervé JN, Ndom P,. Role of health promotion in cancer control in Cameroon and its utilization by the National Cancer Control Program (NCCP), strategy from 2004 – 2019. Int Res J Public and Environ Health. 2021;8(1):1–7. 10.15739/irjpeh.21.001

13. Moyer VA. Screening for cervical cancer: U.S. Preventive Services Task Force recommendation statement. Ann Intern Med. 2012;156(12):880–91, w312. 10.7326/0003-4819-156-12-201206190-00424

14. Mwenda V, Mburu W, Bor JP, Nyangasi M, Arbyn M, Weyers S, et al. Cervical cancer programme, Kenya, 2011-2020: lessons to guide elimination as a public health problem. Ecancermedicalscience. 2022;16:1442. 10.3332/ecancer.2022.1442 PMC9470178

15. Ng’ang’a A, Nyangasi M, Nkonge NG, Gathitu E, Kibachio J, Gichangi P, et al. Predictors of cervical cancer screening among Kenyan women: results of a nested case-control study in a nationally representative survey. BMC Public Health. 2018;18(Suppl 3):1221. 10.1186/s12889-018-6054-9 PMC6219012

16. Ministry of Health Kenya. Kenya STEPwise Survey for non communicable diseases risk factors 2015 report 2015 [Available from: https://www.nutritionhealth.or.ke/wp-content/uploads/Downloads/Kenya%20STEPwise%20Survey%20for%20Non-Communicable%20Diseases%20Risk%20Factors%20Report%202015.pdf.

17. Gebreegziabher ZA, Semagn BE, Kifelew Y, Abebaw WA, Tilahun WM. Cervical cancer screening and its associated factors among women of reproductive age in Kenya: further analysis of Kenyan demographic and health survey 2022. BMC Public Health. 2024;24(1):741. 10.1186/s12889-024-18148-y

18. Kenya National Bureau of Statistics, Ministry of Health Kenya, National Aids Control Council Kenya, Kenya Medical Research Institute, National Council for Population Development Kenya. Kenya Demographic and Health Survey 2014 Rockville, MD, USA2015 [Available from: http://dhsprogram.com/pubs/pdf/FR308/FR308.pdf.

19. Ministry of Health Kenya. Kenya National Cancer Screening Guidelines Nairobi2018 [Available from: https://arua-ncd.org/wp-content/uploads/2022/10/National-Cancer-Screening-Guidelines-2018.pdf.

20. Ministry of Health Kenya. National guidelines for cancer management in Kenya Nairobi2013 [Available from: https://kehpca.org/wp-content/uploads/2020/06/National-Cancer-Treatment-Guidelines2.pdf.

21. Gatune JW, Nyamongo IK. An ethnographic study of cervical cancer among women in rural Kenya: is there a folk causal model? Int J Gynecol Cancer. 2005;15(6):1049–59. 10.1111/j.1525-1438.2005.00261.x

22. Arifin WN. Introduction to sample size calculation. Education in Medicine Journal. 2013;5(2). 10.5959/eimj.v5i2.130

23. Mickey RM, Greenland S. The impact of confounder selection criteria on effect estimation. Am J Epidemiol. 1989;129(1):125–37. 10.1093/oxfordjournals.aje.a115101

24. Zhang Z. Model building strategy for logistic regression: purposeful selection. Ann Transl Med. 2016;4(6):111. 10.21037/atm.2016.02.15 PMC4828741

25. Desta M, Getaneh T, Yeserah B, Worku Y, Eshete T, Birhanu MY, et al. Cervical cancer screening utilization and predictors among eligible women in Ethiopia: A systematic review and meta-analysis. PLoS One. 2021;16(11):e0259339. 10.1371/journal.pone.0259339 PMC8568159

26. Ducray JF, Kell CM, Basdav J, Haffejee F. Cervical cancer knowledge and screening uptake by marginalized population of women in inner-city Durban, South Africa: Insights into the need for increased health literacy. Womens Health (Lond). 2021;17:17455065211047141. 10.1177/17455065211047141 PMC8474337

27. Omoyeni O, Tsoka-Gwegweni J. Knowledge, attitudes and practices of cervical cancer screening among rural women in KwaZulu-Natal, South Africa. Pan Afr Med J. 2022;42:188. 10.11604/pamj.2022.42.188.26172 PMC9508371

28. World Health Organization. Global strategy to accelerate the elimination of cervical cancer as a public health problem 2020 [Available from: https://www.who.int/publications/i/item/9789240014107.

29. Kenya National Bureau of Statistics. The Kenya poverty report. Based on the 2021 Kenya Continuous Household Survey. 2021. https://www.knbs.or.ke/download/the-kenya-poverty-report-2021/:

30. Rosser JI, Njoroge B, Huchko MJ. Knowledge about cervical cancer screening and perception of risk among women attending outpatient clinics in rural Kenya. Int J Gynaecol Obstet. 2015;128(3):211–5. 10.1016/j.ijgo.2014.09.006 PMC4329271

31. Fentie AM, Tadesse TB, Gebretekle GB. Factors affecting cervical cancer screening uptake, visual inspection with acetic acid positivity and its predictors among women attending cervical cancer screening service in Addis Ababa, Ethiopia. BMC Women’s Health. 2020;20(1):147. 10.1186/s12905-020-01008-3

32. Ministry of Public Health and Sanitation Kenya, Ministry of Medical Services Kenya. National guidelines for prevention and management of cervical, breast and prostate cancers. Nairobi2012. http://guidelines.health.go.ke:8000/media/National_Guidelines_for_Prevention_and_Management_of_Cervical_Breast_and_Prostate_Cancers.pdf:

33. National Cancer Control Program. Cancer prevention and early detection Nairobi2023 [Available from: https://nccp.or.ke/.

34. Zhang M, Sit JWH, Chan DNS, Akingbade O, Chan CWH. Educational Interventions to Promote Cervical Cancer Screening among Rural Populations: A Systematic Review. Int J Environ Res Public Health. 2022;19(11). 10.3390/ijerph19116874 PMC9180749

35. Okyere J, Aboagye RG, Seidu AA, Asare BY, Mwamba B, Ahinkorah BO. Towards a cervical cancer-free future: women’s healthcare decision making and cervical cancer screening uptake in sub-Saharan Africa. BMJ Open. 2022;12(7):e058026. 10.1136/bmjopen-2021-058026 PMC9345091

36. Nega AD, Woldetsadik MA, Gelagay AA. Low uptake of cervical cancer screening among HIV positive women in Gondar University referral hospital, Northwest Ethiopia: cross-sectional study design. BMC Women’s Health. 2018;18(1):87. 10.1186/s12905-018-0579-z

37. Yimer NB, Mohammed MA, Solomon K, Tadese M, Grutzmacher S, Meikena HK, et al. Cervical cancer screening uptake in Sub-Saharan Africa: a systematic review and meta-analysis. Public Health. 2021;195:105–11. 10.1016/j.puhe.2021.04.014

38. Ministry of Health Kenya, National AIDS and STI Control Program. Kenya National guidelines for prevention, management and control of sexually transmitted infections Nairobi: NASCOP; 2018 [Available from: Available from: https://www.studocu.com/row/document/mount-kenya-university/hivaids-and-drug-abuse/final-sti-guidelines-17th-october-2018/36652242.

39. World Health Organization Kenya. On the path to expanding cervical cancer screening in Kenya 2023 [Available from: https://www.afro.who.int/countries/kenya/news/path-expanding-cervical-cancer-screening-kenya.

40. Mose D, Katana G, Chea S, Ayieko P. Data from: Uptake of cervical cancer screening and associated factors among women aged 18-49 years attending care at Malindi sub county hospital in Coastal Kenya. figshare; 2024. https://figshare.com/account/projects/200296/articles/25515388:

